# Accelerated biological age linked to high normal serum sodium in general healthcare electronic medical records and NHANES

**DOI:** 10.64898/2026.08.23.26361167

**Authors:** Jonathan Rabinowitz, Ohad Green, Dayoon Kwon, Nuriel Burak, Mahmoud Darawshi, Daniel W. Belsky

**Affiliations:** Wellness Research Lab, Louis and Gabi Weisfeld School of Social Work, Bar-Ilan University, Ramat Gan, Israel; Department of Behavioral Health Science & Practice, College of Behavioral & Community Sciences, University of South Florida. Tampa, FL, USA; Department of Epidemiology, Stanford University School of Medicine, Stanford, CA, USA; Leumit Health Services, Tel Aviv, Israel; Leumit Health Services, HaShalom Clinic, Nazareth, Israel; Mailman School of Public Health, Columbia University, New York, NY, USA

**Keywords:** Hydration, Biological Age, Sodium, Healthy ageing

## Abstract

Recent epidemiological studies suggest poor hydration is a modifiable risk factor for aging-related chronic disease. We tested whether serum sodium was associated with accelerated biological aging. We analysed data from 363,286 adults (18-80 years) from 20 years of electronic medical records from a large healthcare system, as well as 24,611 adults (18-80 years) from National Health and Nutrition Examination Survey (NHANES) continuous (1999-2018). Seven key biomarkers were used to calculate biological age (BA) using the Klemera and Doubal method. We then reran the calculation using only the four variables with highest correlation with age as a robustness check. In both models, there was a significant linear association between age adjusted serum sodium and advanced biological aging, especially in the young cohorts. In the 7-variable model, in the Leumit dataset, the males in the highest sodium level versus the lowest, had a biological age that was 0.88 (95% CI 0.68- 1.08) years accelerated and for females 2.32 (2.14-2.51) years. In NHANES dataset biological age of males at the highest sodium level was 1.92 (0.98-2.87) years accelerated as compared to those in the lowest sodium group. For females, the largest difference was for those 41-50 (1 year, .30-1.79). Increased serum sodium in the normal range is associated with accelerated biological aging in the general population, especially among people aged 18-50. Intervention studies are needed to confirm the link between hydration and biological aging.

## Introduction

Population aging is driving a global epidemic of age-dependent chronic diseases, including cardiovascular, chronic respiratory, musculoskeletal, neurological, and mental disorders. It is estimated that these disorders in individuals aged 60 years and older account for approximately 23% of the total global disease burden, and this proportion is rapidly increasing as life expectancy rises. Consequently, identifying mechanisms and implementing preventive measures to decelerate the aging process have emerged as new challenges for biomedical research and public health (1).

Recent findings suggest that optimal hydration could serve as a potentially relevant factor in the aging process as it might reduce the risk of chronic diseases and result in lower biological age (BA). BA is an estimation of how old a person’s body is compared to same-age peers, based on biomarkers. This hypothesis about the association between hydration and biological age originates from several long-term longitudinal epidemiological studies in which indicators of poor hydration - such as elevated serum sodium and tonicity and increased plasma vasopressin (AVP) concentrations - have been linked to a heightened risk of various health conditions, including hypertension (2), diabetes, heart failure (2–4), kidney disease (5), dementia (6) as well as premature mortality (4, 7) and accelerated biological aging in middle-age adults (4). Support for this hypothesis comes from an intervention study in mice, in which lifelong water restriction shortened their lifespan and led to degenerative changes that suggest accelerated biological aging (8).

Hydration status is a newly identified but understudied factor in chronic disease risk (9). Emerging data from population-based cohorts and experimental research demonstrate that the associated risks of underhydration begin to increase even at mild, subclinical levels of underhydration, where hydration biomarkers remain within currently accepted reference ranges (9). This is particularly relevant considering that over half of the global population consumes less water than recommended (10, 11). Consequently, encouraging adequate hydration may serve as a cost-effective and broadly applicable approach to reducing the burden of accelerated biological aging.

In this study, we leveraged electronic medical records from Leumit Healthcare Services, one of Israel’s four healthcare providers, as well as the National Health and Nutrition Examination Survey (NHANES) continuous (1999–2018), to explore associations between hydration and biological aging. We used serum sodium as a proxy for hydration.

Serum sodium reflects distinct physiological water balance regulatory mechanisms: sodium is the primary contributor to extracellular osmolality (12). We focused on serum sodium as our proxy hydration marker, due to its widespread testing in general medical practice as part of the basic metabolic panel, making it a globally accessible indicator for identifying at-risk individuals. A positive association of serum sodium with accelerated biological aging has been previously established among middle-age persons in the Atherosclerosis Risk in Communities (ARIC) study (4). In the current study, we aimed to confirm these associations using both data from general healthcare record and cross-sectional survey across the adult age range. We quantified biological aging using the algorithm proposed by Klemera and Doubal (13), following the approach used in the ARIC analysis (4).

## Methods

### Population and dataset

The Leumit dataset was provided by Leumit Healthcare Services (hereafter Leumit), one of four national health funds under universal health care in Israel. Leumit provides medical coverage to 7.5% of the total population of Israel (about 720,000 members, 9.2 million patient visits a year) country wide. They have accumulated over 20 years of comprehensive and fully electronic medical records documenting all patient visits across 330 clinics nationwide. All medical information is stored in a centralized electronic medical record (EMR) database, ensuring that no information is missing or exists outside of this system (https://www.innovation.leumit.co.il/about). In this dataset, sample selection bias is minimized, because it is illegal for healthcare service organizations in Israel to refuse membership based on demographic factors, health conditions, or medication needs.

The NHANES is a US cross-sectional survey. It employs a complex survey design and sampling to assess the population’s health and nutritional status. We chose to analyse data from the NHANES continuous survey, which used the same set of questionnaires for every cycle. We included 10 cycles over 20 years (1999–2018). We used the updated curated dataset of (14), which includes cleaned version of the dataset and is freely available to researchers on Github.

Inclusion and exclusion criteria and cohorts selected for analyses

### The Leumit Dataset

The dataset we requested and received from Leumit included males and females over the age of 18 years, who had at least two serum sodium alongside fasting blood glucose measurements after age of 18 years. The provided dataset excluded individuals who had a diagnosis of major chronic diseases at the time of their first sodium test or were diagnosed within the following 2 years. These diseases included: Heart failure (ICD-9 - 428.x), Dementia (ICD-9 codes - 290.x), Chronic Obstructive Pulmonary Disease (ICD-9 codes - 491.x), Asthma (ICD-9 codes - 493.x), Chronic Pulmonary Heart Disease (ICD-9 codes - 416.x), Cardiac Dysrhythmias ((ICD-9 codes - 427.x), Peripheral Vascular Disease ((ICD-9 codes - 443.x), Diabetes Mellitus (ICD-9 codes - 250.x), Stroke(ICD-9 codes - 436.3), Chronic Kidney Disease (Stages I to V) and Renal Failure (ICD-9 codes - 585.x; 586) and Chronic Liver Disease (ICD-9 codes −517.x). Persons with a diagnosis of HIV at any time were also excluded for privacy reasons. Individuals with a diagnosis of hypertension were excluded at later stages of dataset preparation for final analysis. The final dataset provided by Leumit, after these exclusions, contained 479,548 individuals. We have previously provided a more complete description of study cohort selection (15)).

Since the purpose of this analysis was to examine effects of hydration, we aimed to exclude people whose serum sodium could be affected by other factors in addition to amount of liquids they consume. For this purpose, we used the same exclusion criteria as in previous studies (4). Thus, to avoid including people with possible abnormalities of water/salt balance regulation, we additionally excluded people who had average sodium concentration from first two tests outside normal reference range of 135-145 mmol/L and those who were diagnosed with diabetes by the time of second sodium test. Consistent with our previous analysis of this dataset (2), we also excluded individuals who were already diagnosed with hypertension (ICD-9 codes 401.xxx) or HF (ICD9 - 428.x) by the time of second sodium measurement, which was used as baseline time point. After these exclusions, 407,187 individuals remained. Biological age could be calculated for 363,286 of these individuals, who constitute the analytic sample.

### The NHANES Dataset

We endeavoured to apply the same inclusion criteria as in the Leumit dataset, with a few exceptions. First, participants with a single serum sodium test were included as NHANES only measures biomarkers once. Second, as NHANES did not include a direct question about kidney or renal disease (rather than ‘’Renal/Kidney diet’’), we calculated eGFR using the CKD-EPI 2021 formula (16) and excluded those with values below 60. Third, as NHANES is a nationwide survey, all health-related questions are based on self-recall (“Have you ever been told you have…”) rather than recorded medical conditions. After these exclusions, 24,611 individuals remained in the final dataset.

### Exposure variables

Serum sodium was the exposure variable used as a proxy for hydration status, as it increases when body water decreases, which mediates the release of antidiuretic hormone (ADH) and activates water preservation mechanisms (17–19).

#### Serum sodium

For the electronic medical record data, the exposure variable was the average serum sodium derived from the first two blood tests for each individual. This was done to minimize random fluctuations of test results or test day hydration. Similar to previous studies, we conducted the analysis under the assumption that the average of these two serum sodium measurements reflects each individual’s hydration habits (2, 4, 20). The NHANES dataset only included one serum sodium measurement.

### Calculation of biological age (BA)

We calculated BA using the Klemera and Doubal method (13). BA estimated by this method is a superior predictor of mortality relative to chronological age (21) and already shows meaningful variation in young adults (22). We selected seven biomarkers for BA calculation based on knowledge about their age dependency, role in aging process, good performance in previous BA calculations ((13, 21, 23–26)), their availability in most of electronic medical records, correlation with age, and coverage of multiple organ systems and processes: cardiovascular (systolic blood pressure, MCV), renal (urea nitrogen, creatinine), metabolic (cholesterol), Glycaemic (fasting glucose in Leumit and the more stable glycohemoglobin in NHANES) and immune/inflammatory (albumin). The first round of analysis included all seven variables. A second KDM model was fit using only the four biomarkers most strongly correlated with age across both datasets and sexes: blood pressure, glucose/ glycohemoglobin, blood urea nitrogen (BUN), and cholesterol. This excluded variables in the seven-biomarker model that showed weak age correlations or large between-dataset correlation gaps, which can inflate unexplained variance and reduce KDM precision, particularly in NHANES given its survey design and weighting (27).

To calculate BA from selected biomarkers, we used the Klemera and Doubal method (KDM) using the package for R developed by Kwon and Belsky (26). Prior to analysis, we Winzorized the data sets to limit the influence of extreme outliers by setting values below the 5th percentile to the 5th percentile and values above the 95th percentile to the 95th percentile. The data set was randomly split into a test (50%) and modelling (50%) dataset and analysis using KDM were conducted separately for males and females. Missing biomarker values were not imputed. The Bioage Package calculated values for individuals with at least four biomarkers and prorated biological age estimates to account for missing values.

Data were analysed using General Linear Model Univariate procedure that provides regression analysis and analysis of variance for one dependent variable by one or more factors and/or variables. The dependent variable was KDM advance (the difference between an individual’s chronological age and their estimated biological age), sodium level was included as a fixed factor and age as a covariate. An additional round of analysis was conducted to examine the relationship between sodium levels and biological aging within age decade cohorts. Sensitivity analysis was conducted by repeating the analysis controlling for BMI measured within 2 years of baseline which was only available for a subset of medical record data. An additional sensitivity analysis was conducted using a limited set of biomarkers excluding those which could be sensitive to hydration, albumin increases with hemoconcentration, and urea/creatinine levels which rise when fluid volume is low. For the NHANES, data was analysed using the complex survey design (SDMVSTRA, SDMVPSU) and serum-sodium subsample weights using the serum sodium specific weight (WT_LBXSNASI) scaled to pooled cycles per CDC guidelines (CDC, n.d). Analyses were conducted in R (version 4.3.1) using the Bioage release version package <u>GitHub -</u> <u>dayoonkwon/BioAge: Biological Age Calculations Using Several Biomarker Algorithms</u>) and SPSS version 31.

### Ethics

Ethical approval to get access to the deidentified dataset and conduct the current study based on preplanned documented analysis was granted by the Leumit Helsinki Institutional Review Board after evaluation and approval of submitted protocol for the analysis reported in this study (protocol LEU-0009-23).

### Role of the funding source

The funders had no role in the study design, data collection, data analyses, interpretation, or writing of the report.

## Results

After randomly splitting the data into modelling and test data sets, biological aging results were available for 77,446 males and 104,197 females in the test data set for Leumit and 12,453 males and 12,158 females in the NHANES dataset. Table 1 presents the background variables of the subjects included in the test data set by sex. Table 2 presents a Pearson correlation matrix of lab test results and chronological age.

**Table 1.** Descriptive statistics of test data sets (mean and standard deviation)

| Variable | Females<br>(LEUMIT) | Females<br>(NHANES) | Males<br>(LEUMIT) | Males<br>(NHANES) |
| --- | --- | --- | --- | --- |
| Age | 36.44 (14.7)<br>n=103,743 | 40.24 (16.63) n =<br>12,768 | 39.4 (14.2)<br>n=76,994 | 40.33 (17.10)<br>n = 13,196 |
| Total Cholesterol | 186.94 (39.6)<br>n=84,474 | 191.84 (35.50) n =<br>12,760 | 188.03 (40.38)<br>n=68,665 | 192.15 (36.60)<br>n = 13,187 |
| UREA-B | 25.39 (7.8)<br>n=101,886 | 24.25 (7.09)<br>n=12,766 | 30.93 (7.86)<br>n=76,078 | 27.89 (7.60)<br>n=13,196 |
| Glycohemoglobin | N/A | 5.33 (0.34)<br>n = 12,748 | N/A | 5.37 (0.35)<br>n = 13,172 |
| Glucose | 88.39 (10.37)<br>n=95,826 | N/A | 91.76 (11.08)<br>n=76,538 | N/A |
| MCV | 84.9 (9.19)<br>n=95,826 | 88.65 (5.06)<br>n = 12,755 | 86.19 (4.28)<br>n=76,230 | 89.77 (4.08)<br>n = 13,169 |
| Systolic BP | 114.55 (15.8)<br>n=101,707 | 115.16 (13.77)<br>n=12,206 | 121.33 (11.08)<br>n=75,470 | 120.67 (11.81)<br>n=12,752 |
| Albumin | 4.27 (0.32)<br>n=63,741 | 4.22 (.030)<br>n=12,768 | 4.45 (0.30)<br>n=71,996 | 4.43 (0.31)<br>n = 13,196 |
| Creatinine | 0.68 (.13)<br>n=102,446 | 0.71 (0.12)<br>n=12,768 | 0.91 (0.15)<br>n=71,446 | 0.94 (0.14)<br>n = 13,196 |
| Sodium | 139.84 (1.90)<br>n=104,197 | 139.20 (1.96)<br>n=12,768 | 140.57 (1.82)<br>n=77,446 | 139.58 (1.93)<br>n = 13,196 |

**Table 2.** Pearson correlations of test results and chronological age in test data sets.

|  | **Females** |  | **Males** |  |
| --- | --- | --- | --- | --- |
|  | **Leumit**<br>**(n=103,743)** | **NHANES**<br>**(n= 12,158)** | **Leumit**<br>**(n=76,994)** | **NHANES**<br>**(n=12,453)** |
| **Total cholesterol** | .47 | .46 | .39 | .32 |
| **Urea B (BUN)** | .49 | .34 | .35 | .23 |
| **Glucose (Leumit)/**<br>**glycohemoglobin**<br>**(NHANES)** | .29 | .41 | .27 | .37 |
| **MCV** | .14 | .19 | .15 | .24 |
| **Systolic blood**<br>**pressure** | .32 | .53 | .25 | .35 |
| **Albumin** | -.21 | -.10 | -.40 | -.39 |
| **Creatinine** | .32 | .13 | .22 | .06 |
| **Sodium** | .25 | .17 | .03 | .01 |

Age-adjusted linear association between serum sodium (mmol/L) and KDM advance (years), overall and by age decade, stratified by sex are presented in Tables 3 and 4. The β (yrs/mmol/L) directly answers the key question in interpretable units. The final column (**Δ145 – 135)** provides a concrete “10 mmol/L contrast”. In both the four and seven variable models in both datasets there was a significant association between higher sodium and advanced biological aging.

**Table 3:**
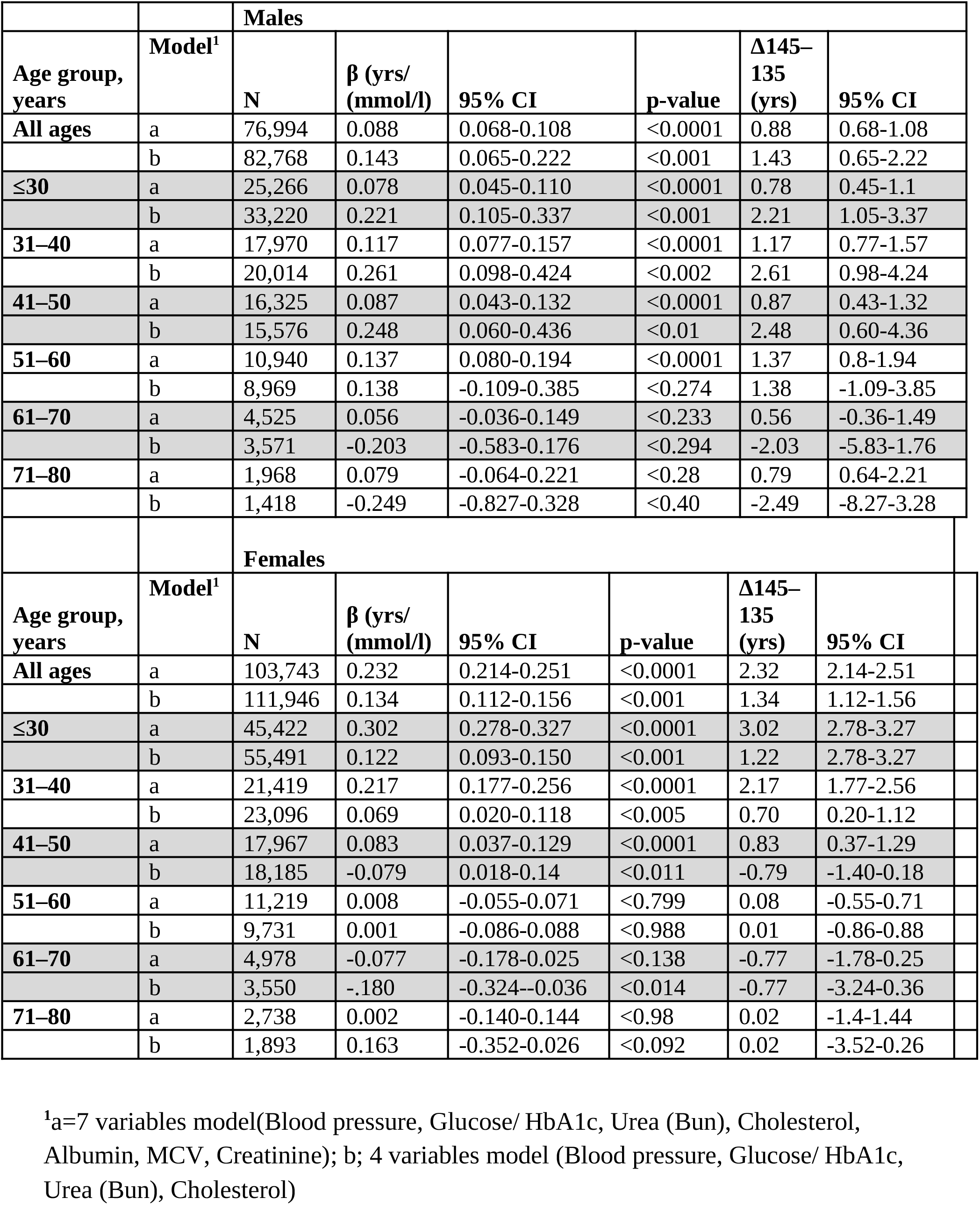
Age-adjusted linear association between serum sodium (mmol/L) and KDM advance (years), overall and by age decade, stratified by sex - LEUMIT dataset.

| Males |  |  |  |  |  |  |  |
| --- | --- | --- | --- | --- | --- | --- | --- |
| Age group, years | Model<sup>1</sup> | N | $\beta$ (yrs/ (mmol/l)) | 95% CI | p-value | $\Delta 145-135$ (yrs) | 95% CI |
| All ages | a | 76,994 | 0.088 | 0.068-0.108 | <0.0001 | 0.88 | 0.68-1.08 |
|  | b | 82,768 | 0.143 | 0.065-0.222 | <0.001 | 1.43 | 0.65-2.22 |
| ≤30 | a | 25,266 | 0.078 | 0.045-0.110 | <0.0001 | 0.78 | 0.45-1.1 |
|  | b | 33,220 | 0.221 | 0.105-0.337 | <0.001 | 2.21 | 1.05-3.37 |
| 31–40 | a | 17,970 | 0.117 | 0.077-0.157 | <0.0001 | 1.17 | 0.77-1.57 |
|  | b | 20,014 | 0.261 | 0.098-0.424 | <0.002 | 2.61 | 0.98-4.24 |
| 41–50 | a | 16,325 | 0.087 | 0.043-0.132 | <0.0001 | 0.87 | 0.43-1.32 |
|  | b | 15,576 | 0.248 | 0.060-0.436 | <0.01 | 2.48 | 0.60-4.36 |
| 51–60 | a | 10,940 | 0.137 | 0.080-0.194 | <0.0001 | 1.37 | 0.8-1.94 |
|  | b | 8,969 | 0.138 | -0.109-0.385 | <0.274 | 1.38 | -1.09-3.85 |
| 61–70 | a | 4,525 | 0.056 | -0.036-0.149 | <0.233 | 0.56 | -0.36-1.49 |
|  | b | 3,571 | -0.203 | -0.583-0.176 | <0.294 | -2.03 | -5.83-1.76 |
| 71–80 | a | 1,968 | 0.079 | -0.064-0.221 | <0.28 | 0.79 | 0.64-2.21 |
|  | b | 1,418 | -0.249 | -0.827-0.328 | <0.40 | -2.49 | -8.27-3.28 |
| Females |  |  |  |  |  |  |  |
| Age group, years | Model<sup>1</sup> | N | $\beta$ (yrs/ (mmol/l)) | 95% CI | p-value | $\Delta 145-135$ (yrs) | 95% CI |
| All ages | a | 103,743 | 0.232 | 0.214-0.251 | <0.0001 | 2.32 | 2.14-2.51 |
|  | b | 111,946 | 0.134 | 0.112-0.156 | <0.001 | 1.34 | 1.12-1.56 |
| ≤30 | a | 45,422 | 0.302 | 0.278-0.327 | <0.0001 | 3.02 | 2.78-3.27 |
|  | b | 55,491 | 0.122 | 0.093-0.150 | <0.001 | 1.22 | 2.78-3.27 |
| 31–40 | a | 21,419 | 0.217 | 0.177-0.256 | <0.0001 | 2.17 | 1.77-2.56 |
|  | b | 23,096 | 0.069 | 0.020-0.118 | <0.005 | 0.70 | 0.20-1.12 |
| 41–50 | a | 17,967 | 0.083 | 0.037-0.129 | <0.0001 | 0.83 | 0.37-1.29 |
|  | b | 18,185 | -0.079 | 0.018-0.14 | <0.011 | -0.79 | -1.40-0.18 |
| 51–60 | a | 11,219 | 0.008 | -0.055-0.071 | <0.799 | 0.08 | -0.55-0.71 |
|  | b | 9,731 | 0.001 | -0.086-0.088 | <0.988 | 0.01 | -0.86-0.88 |
| 61–70 | a | 4,978 | -0.077 | -0.178-0.025 | <0.138 | -0.77 | -1.78-0.25 |
|  | b | 3,550 | -.180 | -0.324--0.036 | <0.014 | -0.77 | -3.24-0.36 |
| 71–80 | a | 2,738 | 0.002 | -0.140-0.144 | <0.98 | 0.02 | -1.4-1.44 |
|  | b | 1,893 | 0.163 | -0.352-0.026 | <0.092 | 0.02 | -3.52-0.26 |

<sup>1</sup>a=7 variables model(Blood pressure, Glucose/ HbA1c, Urea (Bun), Cholesterol, Albumin, MCV, Creatinine); b; 4 variables model (Blood pressure, Glucose/ HbA1c, Urea (Bun), Cholesterol)

**Table 4.** : Age-adjusted linear association between serum sodium (mmol/L) and KDM advance (years), overall and by age decade, stratified by sex - NHANES.

|  |  | **Males** |  |  |  |  |  |
| --- | --- | --- | --- | --- | --- | --- | --- |
| **Age group, years** | **Model<sup>1</sup>** | **N** | **β (yrs/ (mmol/l))** | **95% CI** | **p-value** | **Δ145–135 (yrs)** | **95% CI** |
| **All ages** | a | 12453 | 0.193 | 0.098 to 0.287 | <0.001 | 1.927 | 0.98 to 2.87 |
|  | b | 12453 | 0.286 | 0.180 to 0.392 | <0.001 | 2.86 | 1.80 to 3.92 |
| **≤30** | a | 4623 | 0.284 | 0.155 to 0.412 | <0.001 | 2.836 | 1.55 to 4.12 |
|  | b | 4623 | 0.424 | 0.262 to 0.587 | <0.001 | 4.244 | 2.62 to 5.87 |
| **31–40** | a | 2630 | 0.255 | 0.089 to 0.421 | 0.003 | 2.551 | 0.89 to 4.21 |
|  | b | 2630 | 0.324 | 0.134 to 0.515 | <0.001 | 3.245 | 1.34 to 5.15 |
| **41–50** | a | 2192 | 0.046 | -0.150 to 0.243 | 0.643 | 0.462 | -1.50 to 2.43 |
|  | b | 2192 | 0.1 | -0.119 to 0.320 | 0.367 | 1.004 | -1.19 to 3.20 |
| **51–60** | a | 1412 | 0.091 | -0.090 to 0.273 | 0.321 | 0.914 | -0.90 to 2.73 |
|  | b | 1412 | 0.217 | -0.077 to 0.511 | 0.146 | 2.173 | -.77 to 5.11 |
| **61–70** | a | 938 | 0.019 | -0.278 to 0.316 | 0.899 | 0.191 | -2.78 to 3.16 |
|  | b | 938 | 0.021 | -0.408 to 0.451 | 0.921 | 0.215 | -4.08 to 4.51 |
| **71–80** | a | 546 | 0.111 | -0.173 to 0.396 | 0.438 | 1.114 | -1.73 to 3.96 |
|  | b | 546 | 0.146 | -0.247 to 0.538 | 0.438 | 1.457 | -2.47 to 5.38 |
|  |  | **Females** |  |  |  |  |  |
| **Age group, years** | **Model<sup>1</sup>** | **N** | **β (yrs/ (mmol/l))** | **95% CI** | **p-value** | **Δ145–135 (yrs)** | **95% CI** |
| **All ages** | a | 12158 | 0.034 | -0.005 to 0.072 | 0.089 | 0.335 | -0.05 to 0.72 |
|  | b | 12158 | 0.128 | 0.022 to 0.234 | 0.019 | 1.279 | .22 to 2.34 |
| **≤30** | a | 4333 | 0.066 | 0.017 to 0.115 | 0.009 | 0.659 | 0.17 to 1.15 |
|  | b | 4333 | 0.207 | 0.073 to 0.342 | 0.003 | 2.072 | .73 to 3.42 |
| **31–40** | a | 2669 | 0.047 | -0.018 to 0.111 | 0.152 | 0.469 | -0.18 to 1.11 |
|  | b | 2669 | 0.196 | 0.017 to 0.374 | 0.032 | 1.959 | .17 to 3.74 |
| **41–50** | a | 2330 | 0.105 | 0.030 to 0.179 | 0.006 | 1.048 | 0.30 to 1.79 |
|  | b | 2330 | 0.34 | 0.122 to 0.557 | 0.002 | 3.396 | 1.22 to 5.57 |
| **51–60** | a | 1356 | 0.073 | -0.035 to 0.181 | 0.186 | 0.729 | -0.35 to 1.81 |
|  | b | 1356 | 0.248 | -0.069 to 0.565 | 0.124 | 2.479 | -.69 to 5.65 |
| **61–70** | a | 905 | -0.065 | -0.175 to 0.045 | 0.246 | -0.649 | -1.75 to 0.45 |
|  | b | 905 | -0.196 | -0.513 to 0.122 | 0.225 | -1.957 | -5.13 to 1.22 |
| **71–80** | a | 456 | -0.006 | -0.156 to 0.143 | 0.932 | -0.065 | -1.56 to 1.43 |
|  | b | 456 | 0.114 | -0.311 to 0.539 | 0.594 | 1.141 | -3.11 to 5.39 |

As can be seen in the Leumit dataset (Table 3) there was a significant increase for males with a range of 0.88 from the lowest to highest sodium groups and for females 2.32 years. For males the largest increase was for those in the 51 to 60 age category (1.37 years, n=10,940). For females the largest difference was for the under 30 group, those with the lowest sodium values as compared to the highest had a biological age that was 3 years younger. As can be seen in NHANES dataset (Table 4) there was a significant increase for males with a range of 1.92 years from the lowest to highest sodium groups. For males the largest increase was for those in the under 30 age category (2.83 years, n=4,623). For females no main effect was found, however significant effects were found for specific age cohorts. The largest difference was in the under age 30 (n=4,333) and in the 41-50 (n=2,330) age groups, in both of those age cohorts, those with the lowest sodium values, as compared to the highest, had a biological age that was approximately 1 years younger.

Two sensitivity analyses were conducted. The first sensitivity analysis controlled for BMI and found results consistent with the primary analysis. For the Leumit cohort, for females (n= 39,117) the linear effect across the sodium levels was F=77.69, p<.0001, and for males (n=29,150) F=3.95, p<.0001, Rs=.66, d=.04. For the NHANES cohort, for females (n=12,158) the linear effect across the sodium levels stayed non-significant (F=1.84, p=0.177), Rs=.06, and for males (n= 12,453) it stayed significant (F=12.31, p<0.001). The second sensitivity analysis used variables with no association with hydration: For the Leumit cohort, for females (n=114,080) the linear effect across the sodium levels was F=5.69, p<.0001, and for males (n=83,155) F=15.63, p<.0001. For the NHANES cohort, for females (n= 12,158) the linear effect across the sodium levels stayed non-significant (F=3.75, p=0.055), and for males (n=12,453) it stayed significant (F=18.56, p<0.001).

## Discussion

Using data passively accumulated within electronic health records, we found that, among healthy adults with serum-sodium values in the normal range, higher sodium values were associated with older biological age. In the 7-variable model, effect-sizes were small to moderate [0.019–0.30 years per mmol/L depending on age group and sex] Associations were stronger among women (in the Leumit Dataset) and for persons younger than age 50 (in both datasets).

The observed associations are striking despite their relatively small magnitudes. The data used to calculate biological age came from electronic health records and were generated for a wide range of clinical purpose and from a health survey. Therefore, processes of aging are likely account for only a small fraction of their overall variation. Within this context, a positive association suggests a substantive underlying relationship between hydration, indexed in serum sodium levels, and biological aging, indexed in the Klemera Doubal method algorithm developed for this type of analysis.

A key strength of our study is the robustness of our design for measuring biological aging from electronic health record data. We relied on an established methodological framework, the Klemera-Doubal method (28). We applied this framework to training data obtained from the US National Health and Nutrition Examination Surveys to form our biological age algorithm. Thus, measurement of biological age was defined completely independently of the Leumit data used to conduct hypothesis testing. A further strength is very large size of the Leumit database (363,286) and the wide age range covered by both datasets (18 to 80). Relatedly, although reliance on electronic health records may limit the precision of our biological age observations, it reduces threats of participation bias, because in Israel (as well as other countries such as the UK) health services are offered for free at the point of use. Use of real-world data also provides immediate proof of concept for translating these observations to clinical practice. This is because serum sodium is almost always part of either a basic metabolic panel (BMP) or comprehensive metabolic panel (CMP) (29). However, there are also limitations related to the nature of the observational dataset, which originates from general medical care records that may be subject to incomplete data. This limitation prevented us from performing fully adjusted analyses on the entire cohort. To address this limitation, we also replicated the analyses in the nationally representative, survey-weighted NHANES continuous cohorts, where effect sizes were smaller, but the overall pattern remained consistent for people aged 50 and below. Despite efforts to harmonize the cohorts, important differences persist-particularly between diagnoses officially recorded in medical records and those based on self-reported conditions, as well as between repeated clinical measurements which reduce measurement errors, compared to the single-time biomarker assessments of NHANES. In addition, unlike the Leumit dataset, the serum sodium measure in NHANES was based on a single measurement, while the Leumit cohort used the average of two measures. As a result, hydration may be measured less precisely in NHANES. However, the similar pattern of the two datasets supports the overall validity of the findings. In addition, the non-significant results for over 50 years old in the NHANES dataset can be explained in two ways. First, smaller age groups in the older cohorts which led to a reduced statistical power to detect the small effects. The stronger effect of the NHANES 4-variable model in both males and females, where we removed the variables with low correlation with age might support this assumption. Second, the different health systems of the US and Israel in areas such as access to healthcare, medications and screening. This substantial difference might not reflect in the younger cohorts as they are generally healthier, but only in the older cohorts which are healthy and in need of healthcare services.

Our findings replicate, in an independent electronic-medical-record population and in an NHANES, a continuous, nationally representative survey of the U.S. civilian population the central result reported by Dmitrieva et al (4) that higher-normal serum sodium in midlife - a proxy for habitually lower fluid intake – is associated with biological aging rather than simply with cardiovascular risk.

The consistency of this association across two different cohorts and data sources strengthens the case that the relationship reflects hydration status itself, and not the disease-specific pathways through which sodium is conventionally interpreted. What makes this potentially informative about aging, rather than about sodium per se, is the underlying biology of chronic underhydration. Even mild, subclinical hypohydration that keeps plasma sodium within the normal range is sufficient to switch on water-conservation responses; in animal models, lifelong water restriction raises serum sodium and shortens lifespan, and the proposed mechanism is that the sustained signalling driven by this water-conserving state promotes degenerative changes across multiple organ systems. Viewed this way, serum sodium is of interest here not as a cardiovascular marker but as an accessible readout of a hydration-linked physiological state that may causally modulate the pace of aging.

Because serum sodium testing is inexpensive and near-universally available, it offers a practical screen for individuals. Daily fluid intake guidelines range from 1.6–2.1 L for women and 2–3 L for men (30, 31), yet global surveys indicate over half of people fail to reach even the lower bounds and are frequently underhydrated (10) (11). While interventional studies are still needed to confirm a causal link between hydration and biological aging, individuals with high-normal serum sodium could be flagged for evaluation of their habitual hydration - a candidate modifiable target that, unlike most aging risk factors, is cheap and behaviourally tractable.

## Data Availability

Electronic medical records data provided for analysis by the Leumit Health Services cannot be shared with third parties. NHANES curated dataset is available to download from https://doi.org/10.6084/m9.figshare.21743372

## Acknowledgments

The authors would like to thank the staff of Leumit Health Services and Leumit Start for creating and maintaining electronic medical records and providing access to the data for our analysis.

## Sources of Funding

This research was funded by: Romania’s National Recovery and Resilience Plan (PNRR), Pylon III, Section I8. Development of a Program to Attract Highly Specialised Human Resources from Abroad in Research, Development and Innovation Activities, PNRR-III-C9- 2023-I8, Project “Modelling negative symptom domains neurobiology: a transdiagnostic, translational study”, code CF 46/ 28.07.2023; the Elie Wiesel Chair at Bar-Ilan University, Ramat Gan, Israel, held by the lead author, and by Intramural Research program of the National Heart, Lung, and Blood Institute, NIH, Bethesda, MD, USA: the National Institute of Health grant ZIA-HL006077-10. DWB is a fellow of the CIFAR CBD Network. There was no commercial funding for this work. The funders had no role in study design, data collection, data analyses, interpretation, or writing of this manuscript.

## Statements and Declarations

## Authors’ Contributions

Authorship: JR initiated the study. JR and OG conceptualized the study, wrote and edited the original draft. JR conducted the statistical analysis of the Leumit data and OG the NHANES data. DWB and DK assisted in applying KDM. JR was also involved in funding acquisition and project administration. MD contributed to conceptualizing the study, validating the data and project administration. NB curated and validated the data, managed resources, advised on aspects of methodology relating to understanding the Leumit data. All gave final approval and agreed to be accountable for all aspects of work ensuring integrity and accuracy. JR and OG had direct access to the data.

## Declaration of interest

The authors declare that there is no conflict of interest.

## Notes

### Competing Interest Statement

The authors have declared no competing interest.

### Author Declarations

Leumit Helsinki Institutional Review Board after evaluation and approval of submitted protocol for the analysis reported in this study (protocol LEU-0009-23)

